# Psychological Resilience as a Mediator Between Depression and Quality of Life in Relapsing-Remitting Multiple Sclerosis Patients

**DOI:** 10.1101/2024.06.23.24309357

**Authors:** Yunier Broche-Pérez, Rodneys M. Jiménez-Morales

**Author notes:** Corresponding author information: Yunier Broche-Pérez, PhD., Prisma Behavioral Center, 1696 S Military Trail, Unit C, West Palm Beach, Florida, USA.

## Abstract

Depression represents a significant and prevalent challenge among individuals with multiple sclerosis (MS) substantially impacting their quality of life (QoL). This study explores the mediating role of psychological resilience in the relationship between depression and QoL in a sample of patients with multiple sclerosis (PwMS). This online cross-sectional study involves 179 Relapsing-Remitting Multiple Sclerosis (RRMS) patients. The PwMS completed three questionnaires: the Chicago Multiscale Depression Inventory, the Connor-Davidson Resilience Scale, and the Multiple Sclerosis Quality of Life (MSQOL-29). The results confirmed that higher levels of depression were associated with lower QoL in RRMS patients. However, the inclusion of psychological resilience as a mediator attenuated this direct effect, suggesting that resilience plays a crucial role in mitigating the negative impact of depression on QoL.

## Introduction

Depression represents a significant and prevalent challenge among individuals with multiple sclerosis (MS), substantially impacting their overall well-being and disease management. Studies indicate that between 25% to 50% of patients with MS experience depression during their lifetime, manifesting with a spectrum of symptoms including poor appetite, cognitive difficulties, irritability, discouragement, insomnia, and fatigue (Corallo et al., 2019). Notably, the prevalence of depression varies across different forms of MS, with rates reported at 27.01% for MS overall, 15.78% for relapsing-remitting MS (RRMS), and 19.13% for progressive MS (PMS) (Boeschoten et al., 2017; Feinstein, Magalhaes, Richard, Audet, & Moore, 2014; Wood et al., 2013). Importantly, depression is not merely a psychological burden but also a determinant of quality of life (QoL), affecting treatment adherence and clinical outcomes such as hospital admissions and relapses (Asadollahzadeh et al., 2024; Pilotto, Floris, Solla, Pugliatti, & Zarbo, 2024; Yalachkov et al., 2019). Furthermore, individuals with MS face a significantly heightened risk of suicidal ideation, ranging from 2.3 to 14 times higher than that of the general population, with estimated rates of suicide comprising 1.8% to 15.1% of all MS-related deaths (Kalb, Feinstein, Rohrig, Sankary, & Willis, 2019; Lewis et al., 2017).

Recognizing the profound impact of depression on both psychological and physical health, early diagnosis and effective management are crucial not only for improving quality of life but also for mitigating the risk of suicide in this vulnerable population. Amidst these complexities, the concept of psychological resilience emerges as a pivotal factor that may mediate the relationship between depression and QoL in individuals with MS. Psychological resilience refers to the ability to adapt positively to adversity, encompassing traits such as optimism, adaptive coping mechanisms, and the capacity to maintain psychological well-being in the face of stressors (Broche-Pérez, Jiménez-Morales, Monasterio-Ramos, & Bauer, 2022). Studies suggest that higher levels of resilience are associated with better QoL outcomes across chronic illness populations, including MS, by buffering against the negative effects of depression and other stress-related symptoms (Black & Dorstyn, 2015; Broche-Perez, Jimenez-Morales, Vázquez-Gómez, Bauer, & Fernández-Fleites, 2023; Novak & Lev-Ari, 2023; Ovaska-Stafford, Maltby, & Dale, 2021; Ploughman et al., 2020).

Understanding resilience as a potential mediator between depression and QoL in MS patients holds significant clinical implications. By bolstering resilience through targeted interventions such as cognitive-behavioral therapy (CBT), mindfulness practices, and social support networks, healthcare providers can potentially mitigate the impact of depression on QoL. Effective resilience-building strategies empower individuals to cultivate adaptive coping skills, enhance emotional regulation, and foster a sense of control over their health outcomes, thereby promoting a more positive outlook and improved overall well-being (Forbes & Fikretoglu, 2018; Leppin et al., 2014).

Therefore, this study explores the dynamic interplay between depression, resilience, and QoL in the context of MS, emphasizing resilience as a crucial mediator that may offer pathways to enhance psychological health outcomes and optimize holistic care strategies for individuals navigating the complexities of MS and its associated mental health challenges.

## Methods

Survey data was collected as described in Broche-Perez et al. (2023). In brief, this is an online cross-sectional study involving 179 Relapsing-Remitting Multiple Sclerosis (RRMS) patients. This research is part of the project «Positive modulatory variables of perceived quality of life in patients with multiple sclerosis (MS-POSITIVE project) », coordinated by the Department of Psychology of the Universidad Central «Marta Abreu» de Las Villas, Cuba.

### Measures

#### Chicago Multiscale Depression Inventory CDMI (Sanchis-Segura et al., 2022)

The CMDI is a 42-item, self-report questionnaire that includes three subscales (14 items in each subscale). The three scales are called mood (dysphoria), vegetative symptoms (physical malfunctioning), and evaluative symptoms (negative self-concept and self-criticism). Participants are asked to rate (using a Likert scale from 1 to 5, where 1 is “not at all” and 5 is “extremely”) the extent to which each word describes them during the past week (including today).

#### The Connor-Davidson Resilience Scale 10 items version (CD-RISC 10) (Broche-Pérez et al., 2022)

The CD-RISC 10 is a reduced version of the original 25-item scale (Connor & Davidson, 2003). Patients rate items on a 5-point Likert scale, ranging from 0 (not true at all) to 4 (true nearly all the time). The minimum score that can be obtained in the test is 0 and the maximum is 40. The total score is obtained from the sum of all the items, where higher scores indicate a higher level of resilience.

#### Multiple Sclerosis Quality of Life 29 items version (MSQOL-29) (Rosato et al., 2019)

The MSQOL-29 is a reduced version of the original 54-item inventory (MSQOL-54). This version is made up of seven dimensions of quality of life. The dimensions are ‘bodily pain’, ‘sexual function’; ‘physical function’; ‘emotional well-being’, ‘energy’, ‘cognitive function’, ‘social function’, ‘health distress’, ‘health perceptions’, ‘overall quality of life’, and ‘change in health’. The responses of the patients can be interpreted considering the total score of the inventory and also from two composite scores called *Physical Health Composite*, and *Mental Health Composite*.

### Procedure

All study participants signed an informed consent. The project was approved by the ethics committee of the Department of Psychology of the Universidad Central “Marta Abreu” de Las Villas. Throughout the investigation, the researchers proceeded in accordance with the ethical standards of the 1964 Helsinki Declaration.

### Data Analysis

The data were processed using JASP (JASP, 2024). Descriptive statistics were used to explore the characteristics of the participants. Then, a Pearson’s correlation analysis was performed to assess the correlation between Depression, psychological resilience, and quality of life. To perform a simple mediation analysis, we used bootstrapping sampling (n =5000) distributions to calculate the direct and indirect effects and confidence intervals (95%) of the estimated effects. Significance was determined when the confidence interval does not include zero. Before performing the mediation analysis, lack of multicollinearity, multivariate normality, and linearity were checked. The data was also checked for outliers.

## Results

### Characteristics of the sample

Tables 1 and 2 summarize the demographic and clinical information of the sample (n = 179). The mean age of patients was 40 years. In the sample, 148 participants (82.6%) were females, and most of the participants had a university-level degree (58.6%). Argentina (29.6%), Mexico (15.6%), and Uruguay (14%) were the most represented countries in the study.

**Table 1.**
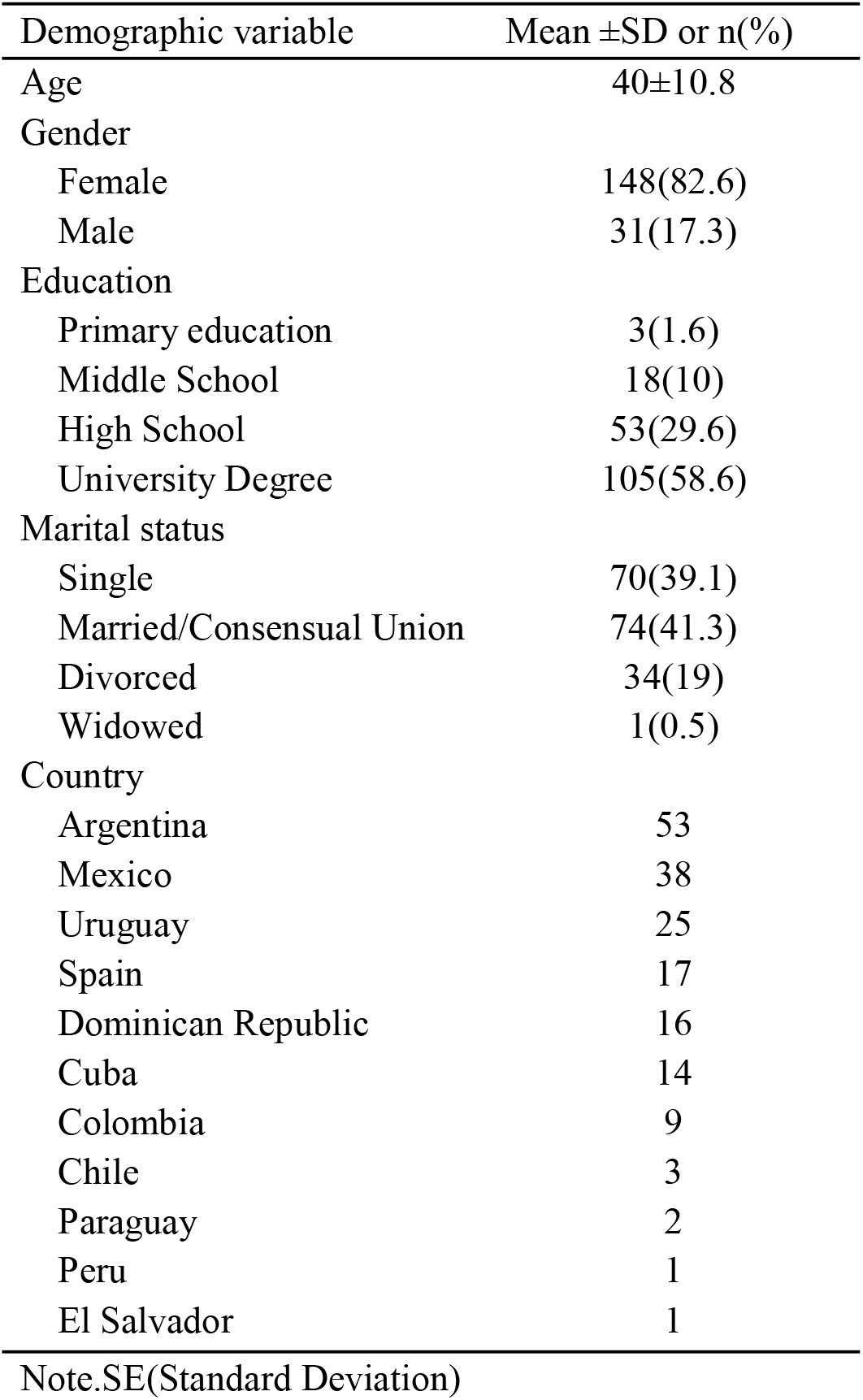
Participant demographic data (n=179)

| Demographic variable | Mean ±SD or n(%) |
| --- | --- |
| Age | 40±10.8 |
| Gender |  |
| Female | 148(82.6) |
| Male | 31(17.3) |
| Education |  |
| Primary education | 3(1.6) |
| Middle School | 18(10) |
| High School | 53(29.6) |
| University Degree | 105(58.6) |
| Marital status |  |
| Single | 70(39.1) |
| Married/Consensual Union | 74(41.3) |
| Divorced | 34(19) |
| Widowed | 1(0.5) |
| Country |  |
| Argentina | 53 |
| Mexico | 38 |
| Uruguay | 25 |
| Spain | 17 |
| Dominican Republic | 16 |
| Cuba | 14 |
| Colombia | 9 |
| Chile | 3 |
| Paraguay | 2 |
| Peru | 1 |
| El Salvador | 1 |

Note. SE(Standard Deviation)

**Table 2.** Participant clinical and psychological data (n=179).

| Clinical variable | Mean±SD or n(%) |
| --- | --- |
| Disease duration<sub>years</sub> | 6.64±7.22 |
| Depression (Total score) | 92.8±29.06 |
| Resilience (Total score) | 26.31±8.34 |
| Quality of Life | 57.9±14.50 |
| Physical Health Composite | 64.21±13.54 |
| Mental Health Composite | 51.98±16.29 |

Note.SE(Standard Deviation)

### Correlation between variables

The analysis of correlations between the variables showed an inverse association between the depression scale scores and the variables resilience and quality of life. As the levels of depression in the PwMS increased, a decrease in the levels of resilience and quality of life was observed. The correlation between resilience and quality of life was positive. As the levels of resilience increase, an increase in the levels of quality of life of patients is also observed (table 3).

**Table 3.** Correlation between variables.

| Variable | 1 |  | 2 |  | 3 |  | 4 |  | 5 |
| --- | --- | --- | --- | --- | --- | --- | --- | --- | --- |
| 1. Depression | — |  |  |  |  |  |  |  |  |
| 2. Resilience | -0.513 | *** | — |  |  |  |  |  |  |
| 3. Quality of life | -0.65 | *** | 0.471 | *** | — |  |  |  |  |
| 4. PHC | -0.422 | *** | 0.281 | *** | 0.806 | *** | — |  |  |
| 5. MHC | -0.719 | *** | 0.519 | *** | 0.85 | *** | 0.467 | *** | — |

\*\*\* p < .001, PHC (Physical Health Composite), MHC (Mental Health Composite)

### Mediation analysis

The mediating effect of resilience between depression and quality of life was explored using a simple mediation analysis (figure 1). As seen in Figure 1, the direct effect of depression on psychological resilience (path a) was significant (β = -.14, 95% CI[-.37, -.26] z = -2.71, p < .001), showing that increasing levels of depression is related to a decrease in resilience levels in our PwMS. Additionally, the direct effect of resilience on quality of life (path b) was also significant (β = .32, 95% CI[.06, .55] z = 2.88, p = .004). When the levels of resilience increase, we can also predict an increase in the QoL in PwMS. On the other hand, path c’ indicates that depression levels predict QoL (β = -.28, 95% CI[.34, .21] z = -8.57, p < .001). According to this result when the levels of depression increase, a decrease in quality-of-life levels can be predicted. The total effect of depression on quality of life was significant (β = -.32, 95% CI[-.37, -.26] z = - 11.44, p < .001), and when psychological resilience, which is the mediator, is included in the model, this effect (β = -.32) decrease (β = -.27), confirming the existence of a significant partial mediation (β = -.04, 95% CI[-.96, .-13], z = 2.71, p = .007). According to this result, psychological resilience acts as a mediator between depression and quality of life in our sample of PwMS.

**Figure 1.**
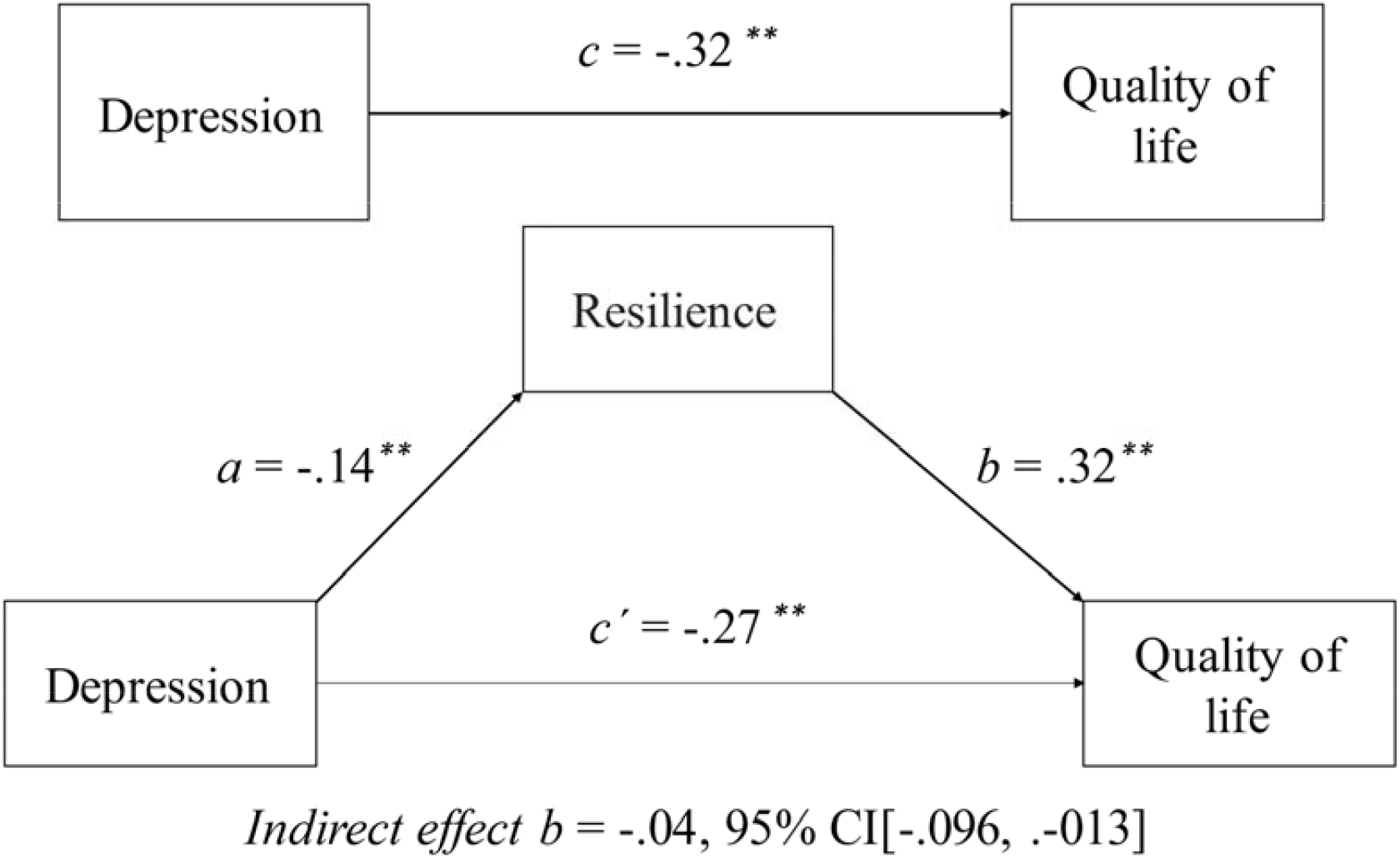
The result of simple mediation model. ^*\*\**^*p* <.001

## Discussion

The present study examined the relationships between depression, psychological resilience, and quality of life (QoL) among individuals with relapsing-remitting Multiple Sclerosis (RRMS). Our findings underscore several important associations and mediation effects that contribute to understanding the impact of depression on QoL in MS and the mediating effect of resilience between depression and QoL.

Consistent with previous research (Broche-Perez et al., 2023; Lee, Rumrill, & Tansey, 2022; Nakazawa et al., 2018; Novak & Lev-Ari, 2023), our results revealed a significant inverse association between depression levels and both resilience and QoL in RRMS patients. As depression severity increased, levels of resilience and QoL decreased. Conversely, a positive correlation was observed between resilience and QoL, indicating that higher resilience levels were associated with better QoL outcomes among RRMS patients. These results highlight that higher levels of resilience are associated with lower levels of depression and higher quality of life scores. This suggests that resilience acts as a protective factor, buffering the negative impact of MS-related symptoms and psychological distress.

On the other hand, a key finding of this study was the mediating effect of psychological resilience between depression and QoL in RRMS patients. Our mediation analysis demonstrated significant pathways: first, the direct effect of depression on psychological resilience was negative and significant, suggesting that higher levels of depression were predictive of lower resilience levels in RRMS patients. Second, the direct effect of resilience on QoL was positive and significant, indicating that higher resilience levels were predictive of better QoL outcomes. Importantly, when considering the total effect of depression on QoL, our findings revealed a significant direct negative effect, confirming that higher levels of depression were associated with lower QoL in RRMS patients. However, the inclusion of psychological resilience as a mediator attenuated this direct effect, suggesting that resilience plays a crucial role in mitigating the negative impact of depression on QoL.

Studies have found that resilience can increase the likelihood of higher quality of life for MS patients, even when risk factors decrease quality of life. One example is the study conducted by Kasser and Zia (2020) aimed to examine the relationship between disease-related risk factors (disability level, fatigue, walking impairments, fear of falling and pain), protective factors (physical activity, self-efficacy, social support, optimism, and health locus of control), coping and resilience on quality of life in adults with multiple sclerosis. According to the results, while higher risk factors decreased quality of life, both directly and indirectly, resilience increased the likelihood of higher quality of life (Kasser & Zia, 2020). Additionally, psychological resilience has also shown a mediating role in the relationship between fear of relapse and quality of life in MS patients (Broche-Perez et al., 2023).

The identification of psychological resilience as a mediator highlights its potential role in interventions aimed at improving QoL outcomes for RRMS patients experiencing depression. Strategies that enhance resilience, such as cognitive-behavioral interventions, mindfulness-based therapies, and social support programs, may offer promising avenues for clinical practice (Bock, Rana, Westemeyer, & Rana, 2024; Giovannetti, Pakenham, et al., 2022; Giovannetti, Solari, & Pakenham, 2022; Pakenham, Mawdsley, Brown, & Burton, 2018). By bolstering resilience, healthcare providers can potentially help RRMS patients better cope with depressive symptoms and improve their overall QoL.

Despite the strengths of this study, including a robust mediation analysis approach, several limitations should be acknowledged. First, the cross-sectional design limits our ability to infer causality between variables. Longitudinal studies are warranted to explore the dynamic interplay between depression, resilience, and QoL over time. Second, the sample consisted of RRMS patients, and findings may not generalize to other MS subtypes or neurological conditions. Future research should consider diverse MS populations to validate these findings across different disease presentations.

## Conclusion

In conclusion, our study advances understanding of the complex relationships between depression, resilience, and QoL in RRMS patients. By elucidating these dynamics and highlighting resilience as a mediator, our findings contribute to the development of targeted interventions aimed at improving psychological well-being and QoL outcomes for individuals living with RRMS. Ultimately, integrating resilience-focused approaches into clinical practice has the potential to enhance patient outcomes and promote holistic care in MS management.

## Data Availability

Data available on request from the authors

